# Future Pandemics: AI-Designed Assays for Detecting Mpox, General and Clade 1b Specific

**DOI:** 10.1101/2024.08.22.24312441

**Authors:** Lucero Mendoza-Maldonado, John MacSharry, Johan Garssen, Aletta D. Kraneveld, Alberto Tonda, Alejandro Lopez-Rincon

## Abstract

The global outbreak of human monkeypox (mpox) in 2022, declared a Public Health Emergency of International Concern by the WHO, has underscored the urgent need for effective diagnostic tools. In August 2024 WHO again declared mpox as a Public Health Emergency of International Concern. This study presents an innovative approach using artificial intelligence (AI) to design primers for the rapid and accurate detection of mpox. Leveraging evolutionary algorithms, we developed primer sets with high specificity and sensitivity, validated in *silico* for mpox main lineage and the Clade 1b. These primers are crucial for distinguishing mpox from other viruses, enabling precise diagnosis and timely public health responses. Our findings highlight the potential of AI-driven methodologies to enhance surveillance, vaccination strategies, and outbreak management, particularly for emerging zoonotic diseases. The emergence of new mpox clades, such as Clade **1b**, with higher mortality rates, further emphasizes the necessity for continuous monitoring and preparedness for future pandemics. This study advocates for the integration of AI in molecular diagnostics to improve public health outcomes.

## Introduction

The global outbreak of human monkeypox, announced by the World Health Organization (WHO) in May 2022, has raised significant public health concerns. Monkeypox virus (mpox), first isolated in 1958 and classified as a species within the *Orthopoxvirus* genus together to the variola virus that causes smallpox, is a zoonotic disease primarily transmitted from animals to humans, with limited human-to-human transmission^1^. The first human case was recorded in 1970 in the Democratic Republic of the Congo. The disease manifests with a distinctive rash, fever, headache, backache, lymphadenopathy, and fatigue, and can lead to severe complications such as sepsis, pneumonia, bacterial super-infection, vision loss, and death^2^. While smallpox vaccines are 85% effective against mpox and a new two-dose vaccine has been approved, the effectiveness of these treatments in the current outbreak is still under study. By June 2022, the outbreak had spread to multiple countries, with over 3,400 confirmed cases and one fatality, predominantly in the WHO European Region and the Region of the Americas^3^.

However, in August 14*th* the WHO Director-General, Dr. Tedros Adhanom Ghebreyesus, declared the recent surge of mpox in the Democratic Republic of the Congo (DRC) and other African countries a Public Health Emergency of International Concern (PHEIC). This decision follows advice from an independent expert committee, which highlighted the potential for the virus to spread further within Africa and further continents. The declaration underscores the rapid spread of a new Clade (**1b**) of mpox in eastern DRC and neighboring countries, necessitating a coordinated international response to control the outbreaks and save lives^4^. Multiple outbreaks of various mpox clades have emerged in several countries, exhibiting diverse transmission methods, primarily through sexual networks, and presenting varying levels of risk^5^. Effective national surveillance, post-exposure vaccination for contacts of cases, pre-exposure vaccination for high-risk groups, and standardized research protocols are crucial to contain the spread of the virus. This outbreak underscores the urgent need for robust public health responses and preparedness for future zoonotic disease outbreaks. Mpox, has two main genetic clades: Clade I and Clade II. Clade I, found mainly in Central and East Africa, is more severe with higher mortality rates. Clade II, which includes subclades IIa and IIb, is generally milder and was responsible for the global outbreak in 2022^6,7^. Recently, a new variant called Clade 1b has emerged. This variant is considered the most dangerous so far, with a higher mortality rate and more severe symptoms^8^.

Recently, the COVID-19 pandemic underscored the essential need for accurate and timely virus detection, particularly for managing the spread of SARS-CoV-2. As new variants of concern (VOCs) with significant mutations emerged, the demand for specific molecular tests capable of precisely identifying these variants increased, given their distinct symptoms. Consequently, with the potential for new pandemics, new software was developed to rapidly create primer sets from a small number of samples to help contain the spread^9,10^. In a previous study, we developed an innovative tool that uses artificial intelligence (AI), specifically evolutionary algorithms (EAs), to design primers for detecting SARS-CoV-2 and its VOCs. By utilizing sequences from the Global Initiative on Sharing All Influenza Data (GISAID) repository^11^, we created an automated pipeline that generates primer sets within hours. These primers were validated both in silico and in laboratory settings, demonstrating high accuracy and superior performance compared to existing commercial versions^10^. This procedure can be used in the same form for mpox.

Primer sets are crucial for detecting mpox because they enable precise identification of the virus’s DNA through polymerase chain reaction (PCR) testing. These sets ensure accurate diagnosis by specifically targeting mpox virus DNA, distinguishing it from other similar viruses. This specificity is vital for rapid detection, allowing for timely treatment and containment of outbreaks. Additionally, primer sets are designed to be highly sensitive, capable of detecting even small amounts of viral DNA, which minimizes the risk of false negatives. Accurate detection also aids in tracking the virus’s spread and understanding its epidemiology, which is essential for effective public health responses^5^. This study highlights the potential of AI-driven approaches in creating efficient and specific primers, offering a promising solution for rapid and accurate detection of viruses and its variants, specifically mpox and its variants, and potentially enhancing responses to future pandemics.

## Results and Discussion

### General Primers

We runt the EA algortihm 20 times, to find suitable forward primers (Fig. 1).

**Figure 1.**
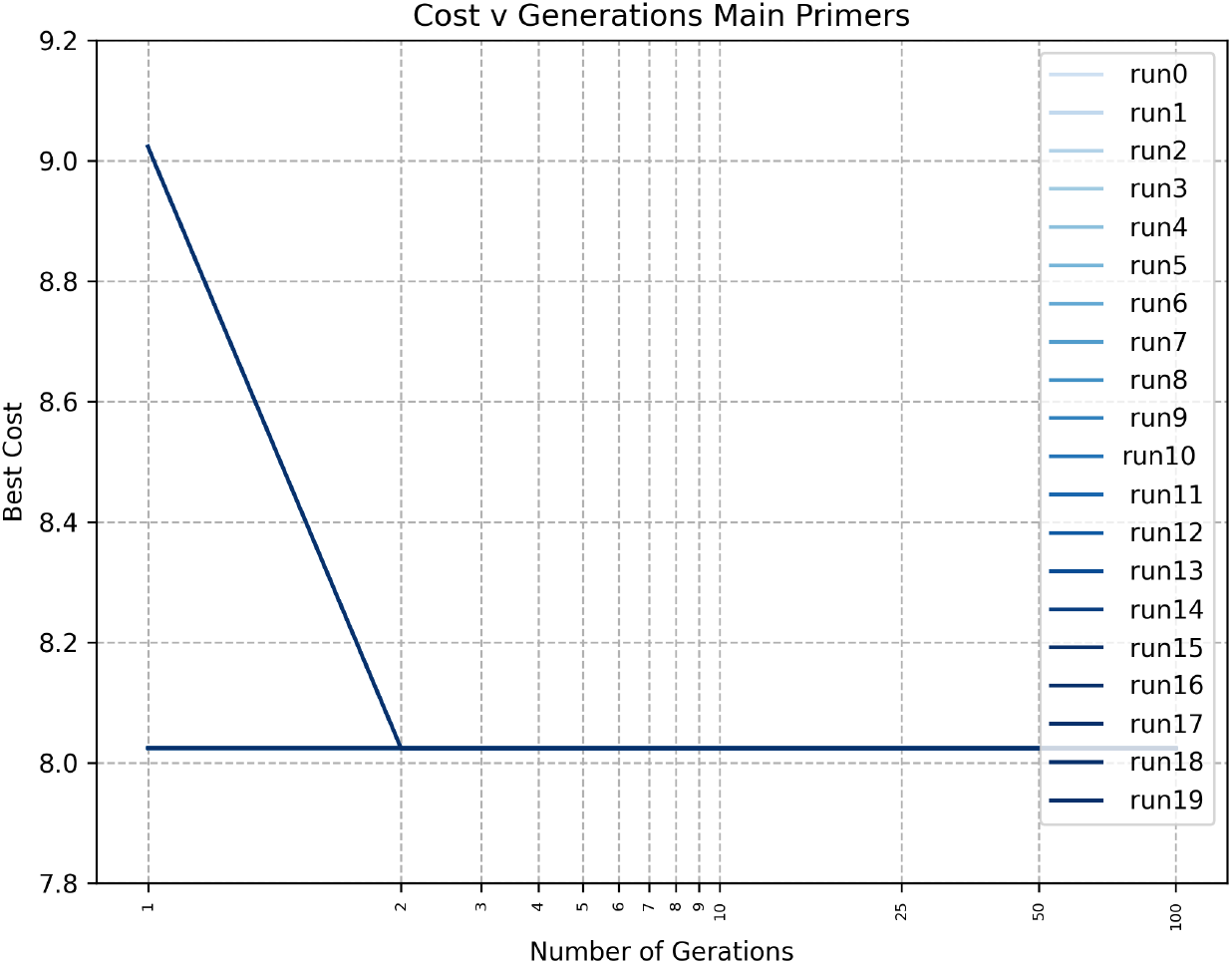
Cost function in 20 runs of the EA for 100 generations to find a forward primer in the main lineage. The shape of the result, is given by the size of the initial population.

We identified four distinct sequences for the forward primer, each demonstrating 100% in-silico specificity and 100% sensitivity. However, according to the analysis in^12^, using the sequence MPXV_USA_2022_MA001^13^ and the reference sequence EPI_ISL_13053218, three of these forward primer sequences are located within the conserved region^12^ (Table 1). In contrast, the sequence **ACGACCTAGACGCCCTACAAA** is outside the conserved region, thus it is susceptible to future mutations.

**Table 1.**
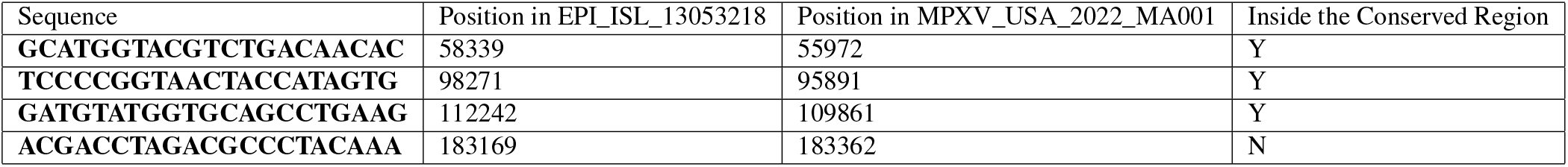
Found Forward Primers for the general primer set.

Next, using Primer3Plus^14^, we generated the internal oligo and reverse primer for each of the forward primers in the conserved region. Finally, we verify the frequency of appearance of the sequences in other taxa (20,603 samples of other viruses) and in mpox (3,515 samples), including the internal oligo and the reverse primer (Table 2).

**Table 2.**
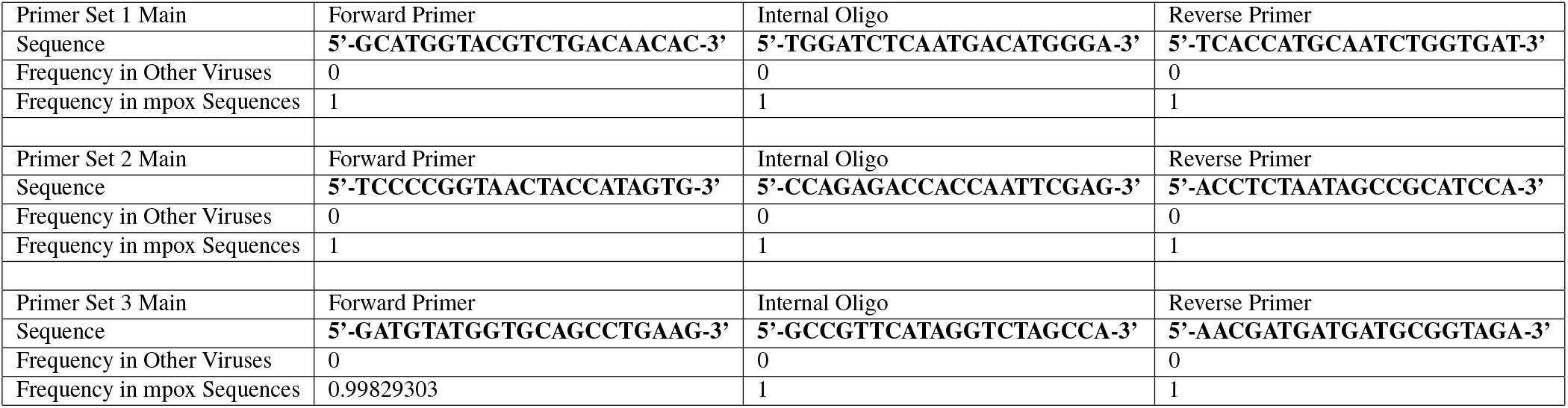
Different options for a general mpox primer set with the frequency of appearance in different taxa sequences (20,603) and mpox sequences (3,515).

### Clade 1b Primers

Again, we ran the EA algorithm 20 times to find suitable specific forward primers for the Clade 1b (Fig. 2).

**Figure 2.**
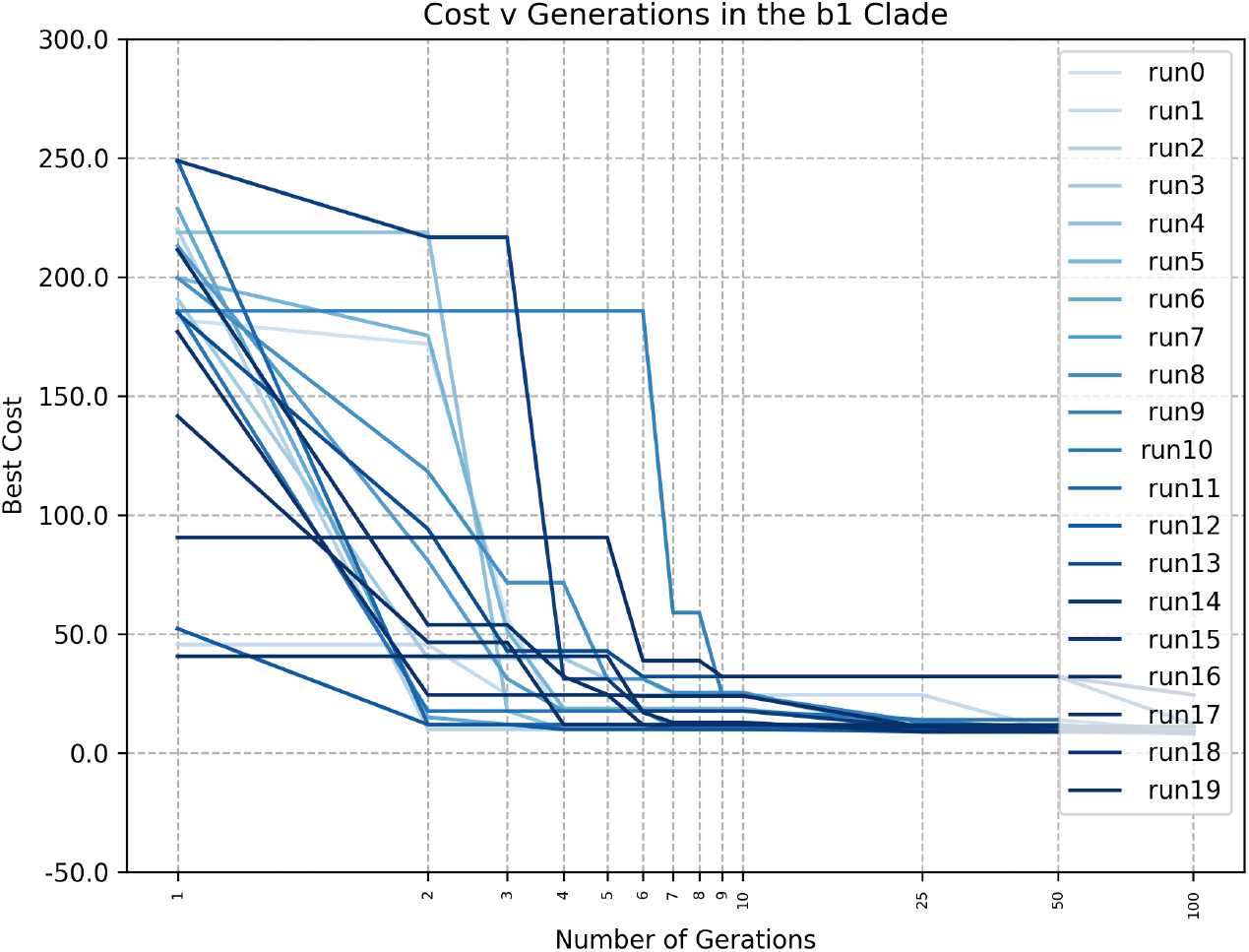
Cost function in 20 runs of the EA for 100 generations to find a forward primer in the main lineage.

We identified six sequences that could be used as forward primer to separate Clade **1b**. However, only one of these sequences had enough mutations to serve as a specific forward primer. Based on the experience described in^10^, a clade/variant primer must contain several mutations to be specific. The sequence, **CGTCGGAACTGTACACCATAG**, is specific for Clade **1b** with a sensitivity of 100% and a specificity of 99.65%. Finally, using Primer3Plus^14^, we then generated the internal oligo and the reverse primer (Table 3).

**Table 3.**
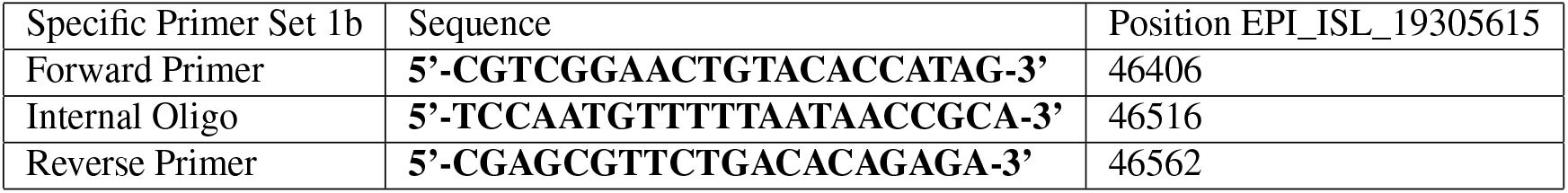
Specific Primer for Clade 1b.

#### Comparison to Other Primer Sets

We made a comparison to other primer sets available in the literature. This include: OPG123^15^, MPXV West African specific (G2R_WA^16^), MPXV Congo Basin specific (C3L^16^) and MPXV generic (G2R_G^16^), sequences available in Table 4.

**Table 4.**
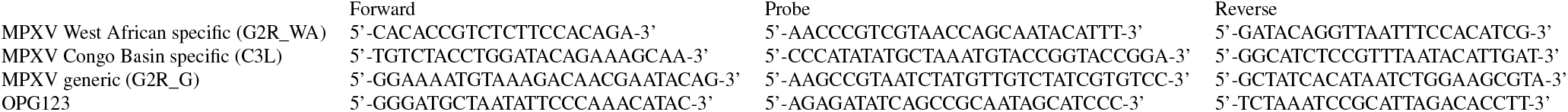
Sequences of other Assays used for mpox.

Using the sequences in Table 4, and ours we get the following results, Fig. 3.

**Figure 3.**
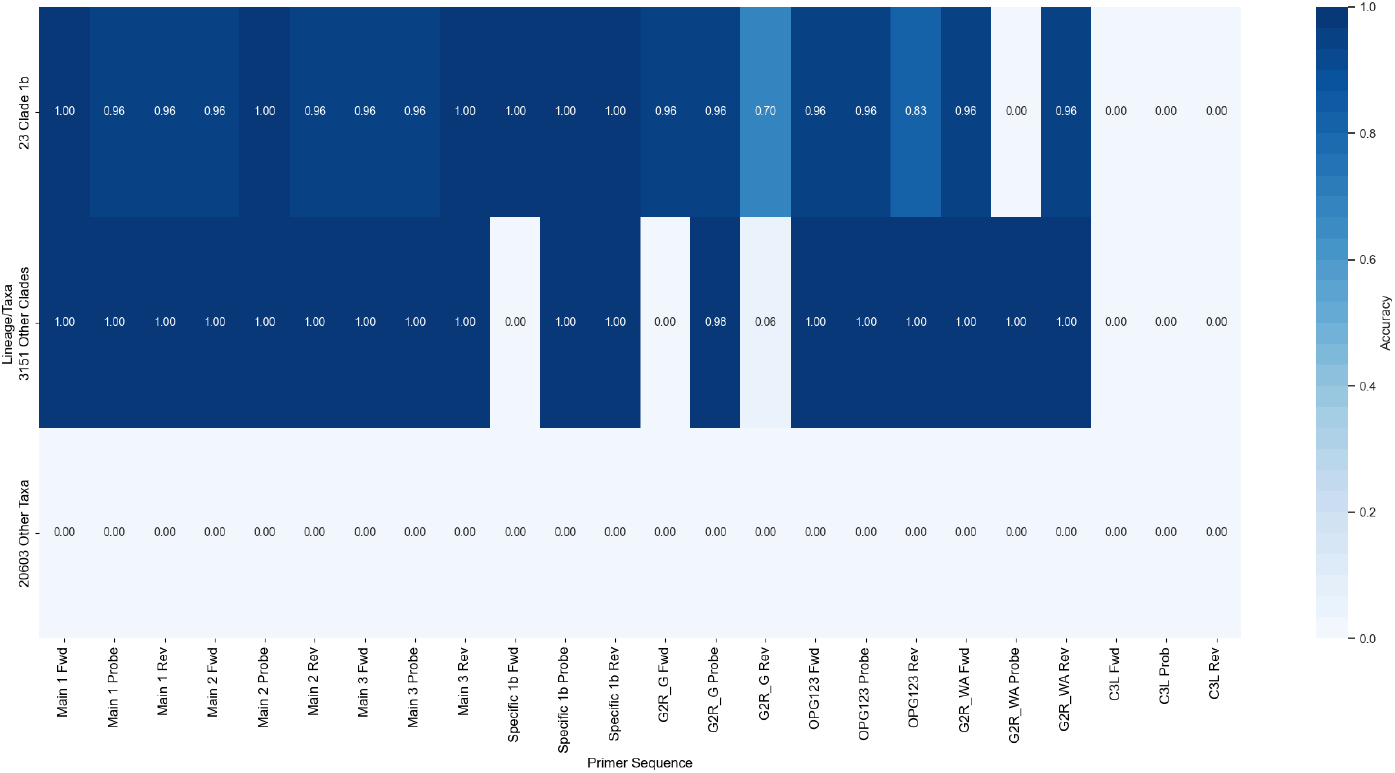
Comparison of different assays, for 3,515 sequences of mpox from GISAID, 23 mpox sequences Clade 1b and 20,603 other taxa.

## Discussion

This study emphasizes the crucial role of AI-driven methods in rapidly and accurately detecting viruses, with a specific focus on mpox. By utilizing evolutionary algorithms, researchers successfully designed primer sets that demonstrated high specificity and sensitivity in detecting mpox and its Clade 1b. This innovative approach, previously validated for SARS-CoV-2 variants of concern (VOCs), highlights AI’s potential to significantly enhance public health responses to emerging zoonotic diseases. Notably, the main assays for mpox (as shown in Table 1) are present in most mpox sequences, including Clade 1b. OPG123 shows similar results, while G2R_G is specific to Clade 1b sequences. In contrast, G2R_WA appears in most sequences but cannot be used for Clade 1b. C3L does not appear in any of the sequences. While our findings are promising, additional validation in controlled laboratory environments remains essential.

Our findings emphasize the need of accurate tools to create fast new diagnostic primers, facilitating timely treatment and containment measures. The study advocates for the implementation of robust surveillance systems, comprehensive vaccination strategies, and standardized research protocols to effectively control the spread of mpox and similar viruses. The emergence of new mpox clades, such as Clade 1b, characterized by higher mortality rates and severe symptoms, further underscores the necessity for continuous monitoring and preparedness for future pandemics.

In conclusion, the integration of AI in primer design presents a promising avenue for improving the accuracy and efficiency of virus detection, ultimately contributing to enhanced public health outcomes. The innovative approach presented in this study can be adapted for other viruses, offering a promising solution for rapid and precise detection in future pandemics.

## Methods and Data

### Data

#### General Primer Set

On March 17*th*, 2020, we downloaded data from NCBI^17^ using the query: “virus” AND host = “homo sapiens” AND “complete genome”, with a size restriction of 1000 to 35,000 bps. This query yielded 20,603 samples from various taxa, including Hepatitis B, Dengue, Human immunodeficiency, Human orthopneumovirus, Enterovirus A, Hepacivirus C, Chikungunya virus, Zaire ebolavirus, Human respirovirus 3, Orthohepevirus A, Norovirus GII, Hepatitis delta virus, Mumps rubulavirus, Enterovirus D, Zika virus, Measles morbillivirus, Enterovirus C, Human T-cell leukemia virus type I, Yellow fever virus, Adeno-associated virus, SARS-CoV-2 and rhinovirus (A, B, and C), totaling over 584 other viruses (excluding strains and isolates). Then, on June 22*nd*, 2022 from the Global Initiative on Sharing Avian Influenza Data (GISAID) repository^11^, we downloaded 191 sequences identified as mpox.

The classification problem was defined as binary classification, with mpox samples labeled with ‘1’, and the rest labeled with ‘0’. For further validation, we downloaded 3,515 sequences from the GISAID repository on August 15*th*, 2024, to verify whether our designed primer set works for the different clades.

#### Clade 1b Primer Set

On August 17*th* 2024, we downloaded 23 sequences mpox Clade **1b** from GISAID repository and 292 from other clades. Again, we assigned “1” to samples to Clade 1b, and “0” to the rest, for a binary classification.

## Methods

### Creating the Primer Set

To create the primer set, we use a technique based on Evolutionary Algorithms (EAs), that is able to find suitable sequences to identify the target samples^18^. In summary, this method optimizes two integer values *p, k* that jointly identify the best candidate forward primer of length 21 at position *p* in sample *k* of the training set, see Figure 4. Then gives a cost based on its frequency of appearance marked with label ‘1’, its absence from samples with label ‘0’ (in other words, its specificity to the target variant), its CG content, and melting temperature, as described in more detail in^10^, where the same technique was used to generate primers VOCs of SARS-CoV-2. The cost function in our study assesses the suitability of a candidate sub-sequence as a primer, aims to be maximized, and is defined as follows:

**Figure 4.**
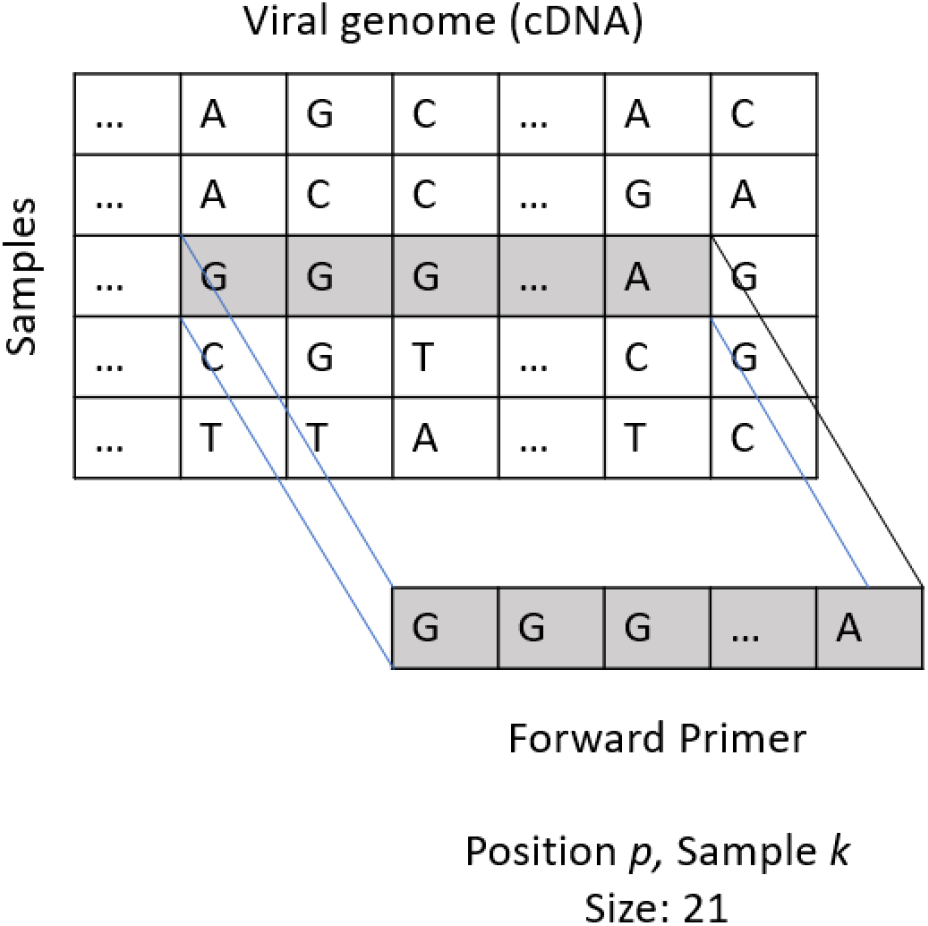
EA system where we select a target 21 bps sequence in position *p* and in sample *k*.

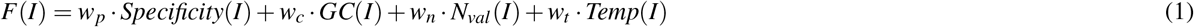

with *w*_*p*_, *w*_*c*_, *w*_*n*_, *w*_*t*_ representing the weights associated to each term.

As EAs are stochastic algorithms, and can thus potentially return different candidates at each run, we run the algorithm 20 times to obtain a variety of candidate sequences, with 200 as initial population, tournament 2 for 100 iterations. Once a candidate sequence is identified, we simulate it in Primer3Plus^14^ using the accession *EPI*_*ISL*_13053218 as a reference for the general primers of mpox and *EPI*_*ISL*_19305615 for the specific primers for Clade **1b**.

The used data for this manuscript, and the resulting frequency of appearance for each sequence is available in: https://github.com/steppenwolf0/primersMPOX2024

## Data Availability

The exact data belongs to GISAID, we shared just the results from the analysis in the Github. The IDs of the viral samples are available in the Github.

https://github.com/steppenwolf0/primersMPOX2024

## Additional information

J. Garssen is a part time employee at Danone Nutricia Research, Utrecht, the Netherlands. No conflict of Interest of any of the authors.

